# Research on the Trend of the Height of Han Students Aged 7-18 in China

**DOI:** 10.1101/2020.06.16.20122994

**Authors:** Chenlu Hong, Zhang Rui, Fei Wu

**Author notes:** These authors contributed equally to this work. These authors also contributed equally to this work.

## Abstract

**Aims:** To make time series analysis on the 29-year (1985−2014) trends in height among Han Students Aged 7-18 in China and predict their future height from the perspective of interdisciplinary studies of demography and physical education.

**Methods:** The data were from the findings of seven cross-sectional surveys from the Chinese National Survey on Students’ Constitution and Health (CNSSCH). The mean, standard error, variance analysis, trend test, F test, T test, development growth and growth rate were used to make a descriptive analysis of height trends. The ARIMA model in the time series analysis was applied to predict the height development of Han students in 2024.

**Results:** The height of students aged 7-18 substantially increased from 1985 to2014. The mean of the height of boys increased by 3.99 cm, 8.01 cm, 8.31 cm, 8.63 cm, 9.81 cm, 11.62 cm, 10.38 cm, 9.22 cm, 7.50 cm, 5.59 cm, 4.51 cm, and 3.79 cm, respectively. The mean of the height of girls increased by 6.66 cm, 7.36 cm, 8.00 cm, 8.84 cm, 9.60 cm, 8.66 cm, 5.57 cm, 4.65 cm, 3.95 cm, 3.32 cm, 2.87 cm and 3.32 cm, respectively. The increasing disparity of the sex differential in the mean height was also observed. From 1985 to 2014, the mean of height difference between boys and girls aged 7-18 had been increased constantly. Their mean height differences were 3.27 cm, 4.03 cm, 4.64 cm, and 5.13 cm, respectively in 1985, 1995, 2005, and 2014. A narrowing of the urban−rural differential in the mean height was observed. In 1985, on average, urban boys aged 7-18 were 4.18cm higher than rural boys in the same age group. And the height differences between urban and rural boys in the same age group were 3.58 cm, 3.18 cm and 2.41 cm, respectively in 1995, 2005 and 2014. According to comparison results, the mean height of urban girls was greater than that of rural girls, and the height difference between urban and rural girls had been constantly narrowed as well. The results of 4 comparisons showed that the height differences between urban and rural girls were 3.68 cm, 3.12 cm, 2.62 cm, and 1.98 cm, respectively.

**Conclusions:** There was a general increase in the height of Chinese Han students aged 7-18 in the past 29 years and difference between sex, rural-urban and age have been observed. In 2024, the height of students will continue to grow.

## Introduction

The Chinese National Survey on Students’ Constitution and Health (CNSSCH) was organized and implemented by the Ministry of Education for 7 times from 1985 to 2014, involving 4 aspects: body shape, physical function, physical quality and physical condition. In the 29 years of surveillance on students’ constitution and health, test indicators of physical functions and health had changed a lot, but the test instruments, methods and technical specifications of the body shape indicators changed little, and the tests had a relatively high degree of standardization. Therefore, the data has good consistency and high research values.

Body shape refers to the shape characteristics of the internal and external parts of the human body and, to a certain extent, reflects the level of physical development, physical function and physical quality of people. It is an important index in the constitution test. And the height is the most representative index in the shape development process [1]. The height of boys and girls is a hot topic in physical education, preventive medicine, hygiene, and pediatrics. In this research, height indexes of Chinese Han students aged 7-18 in the period from 1985 to 2014 were analyzed to discuss their development characteristics and laws, and the AMIRA model was used to predict their height development trend in 2024.Taking height indexes in the constitution test as an entry point, this research tried to use new methods and new ideas to analyze related data of the constitution test and helped to deepen the longitudinal development of constitution and health research in China.

## Methods

### Data sources

The data were obtained from the Chinese National Survey on Students’ Constitution and Health (CNSSCH), which has been conducted in China since 1985. In the seven surveys, there were 471,115 in 1985, 140,633 in 1991, 249,492 in 1995, 216,681 in 2000, 291,604 in 2005, 215,319 in 2010, and 214,359 in 2014, respectively, the ratio of boy: girl or urban: rural was approximately 1:1 in each [2-8].

### Measurements

The mean, standard error, variance analysis, trend test, F test, T test, development growth and growth rate were used to make a descriptive analysis of height development trends. The time series analysis was applied to predicting the height indexes of Han students aged 7-18 in 2024.

Firstly, urban and rural indexes were based to calculate the national indexes. The mean, standard deviation, and sample size as relevant urban and rural indexes were known and based to calculate the mean values of height indexes for the National Han students aged 7-18 in 1985, 2000, and 2014.

Secondly, a variance analysis and trend test were made on the height indexes of nationwide students in China.

Thirdly, pairwise year-based t-tests were made of height indexes of Han students aged 7-18 in 1985, 1995, 2005 and 2014. The t-test method was adopted when the variance was homogenous, while the Satterthwaite t’ test was used when the variance is not homogenous.

Fourthly, dynamic analysis indexes for the height of Han students aged 7-18 in the period from 1985 to 2014 were calculated [2.

#### 1. Growth of the development level

Growth is the difference between the levels of development in two different periods in the time series. The calculation formula for the growth is:

Growth = the development level of the report period - the development level of the base period.

1. The period-by-period growth is the growth calculated by taking the previous period as the base period in the time series, that is, a_1_-a_0_, a_2_-a_0_…,a_n_-a_0_.
2. Cumulative growth is the difference between the level of the report period and the level of a fixed period. Let a_0_ be the fixed period level, and the cumulative growth is: a_1_-a_0_, a_2_-a_0_…,a_n_-a_0_.

#### 2. Development growth rate

The growth rate is a relative index which reveals the growth degree of a phenomenon. It is the ratio of the growth rate to the development level of the base period. The growth rate is divided into the link-relative growth rate and the fixed base growth rate according to different base periods.

Link-relative growth rate = period-by-period growth / the development level of the previous period.

Fixed based growth rate = cumulative growth / the development level of the fixed period

Fifthly, the ARIMA model in the time series was used for predictions.

## Results

### Analysis on the overall height trend

During the 29 years, the height of Chinese Han students aged 7-18 continued to grow (Fig 1), but the difference in the overall mean change of the full-range school age group was not significant (Table 1).

**Table 1.** One-way analysis of variance and trend test of Han students’ height from 1985 to 2014.

|  | Boys |  |  |  | Girls |  |  |  |
| --- | --- | --- | --- | --- | --- | --- | --- | --- |
|  | Variance analysis |  | Trend test |  | Variance analysis |  | Trend test |  |
| Index | F-value | P-value | F-value | P-value | F-value | P-value | F-value | P-value |
| Height | 0.28 | 0.95 | 1.63 | 0.20 | 0.26 | 0.95 | 1.54 | 0.22 |

**Fig 1.**
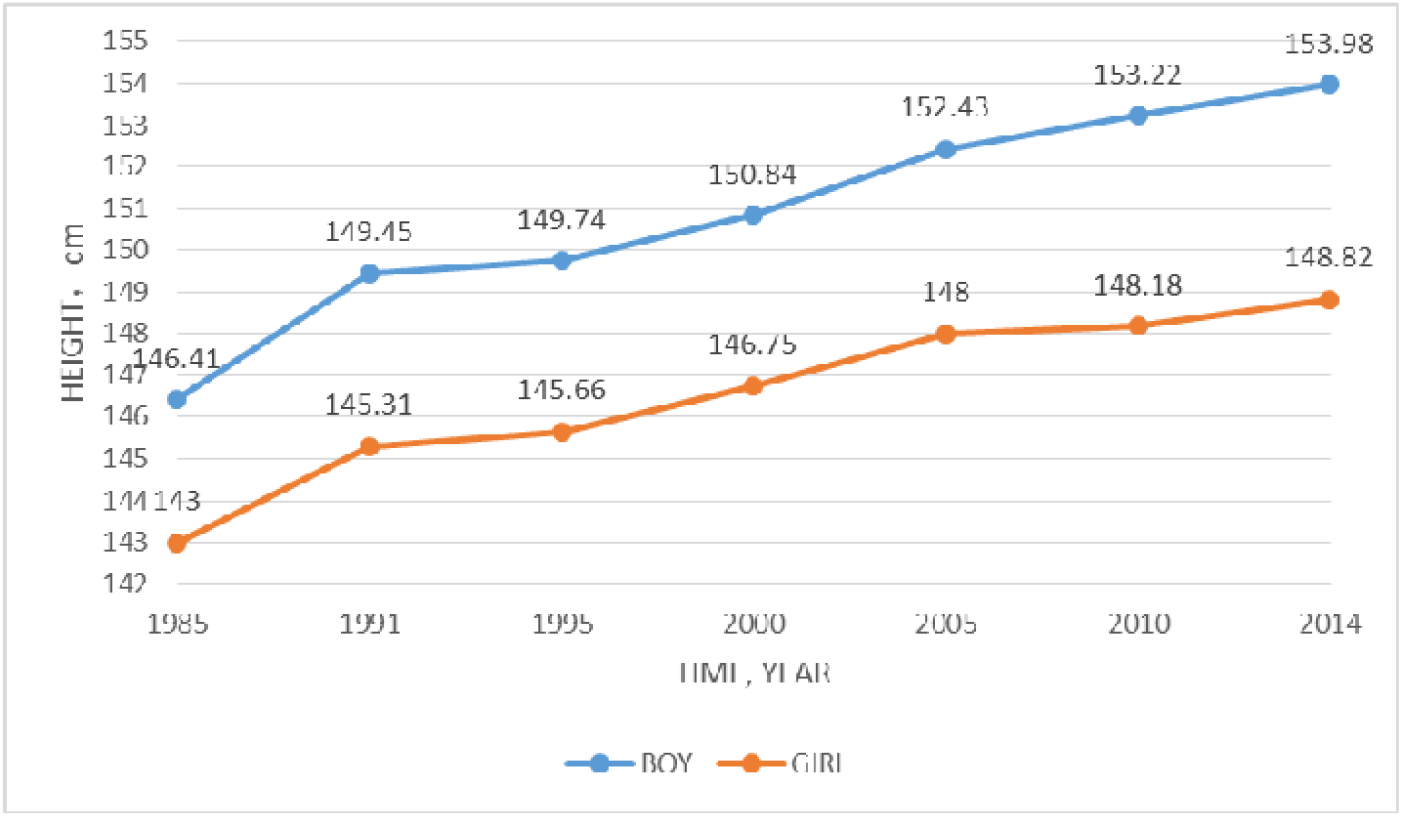
The development trend of the mean height of Han students in the full-range school age group from 1985 to 2014.

### Analysis on the secular height trend

From 1985 to 2014, the mean height of the Han boys aged 7-18 in the same age group showed an upward trend as times went on (the difference is statistically significant, as shown in Table 2 and Table 3). The time series analysis was used to make a dynamic analysis on the cumulative growth of the height indexes. The mean heights of boys aged 7-18 increased by 3.99cm, 8.01 cm, 8.31 cm, 8.63 cm, 9.81 cm, 11.62 cm, 10.38 cm, 9.22 cm, 7.50 cm, 5.59 cm, 4.51 cm and 3.79 cm, respectively. Boys aged 12 had the largest growth (11.62 cm). The mean height of boys in the full-range school age group increased by 7.61cm. The link-relative growth of boys in the full-range school age group was respectively 2.64 cm, 0.65 cm, 0.96 cm, 1.13 cm, 1.26cm, and 1.23cm in the seven tests. According to an analysis on the period-by-period growth, boys had the largest growth from 1985 to 1991. And the height growth of boys aged 7 to 9 in 1991 was respectively 2.32 cm, 2.44 cm, and 2.41 cm, and the corresponding figure of boys aged 12, 13 and 14 was 4.17 cm, 4.32 cm, and 2.62 cm, respectively.

**Table 2.**
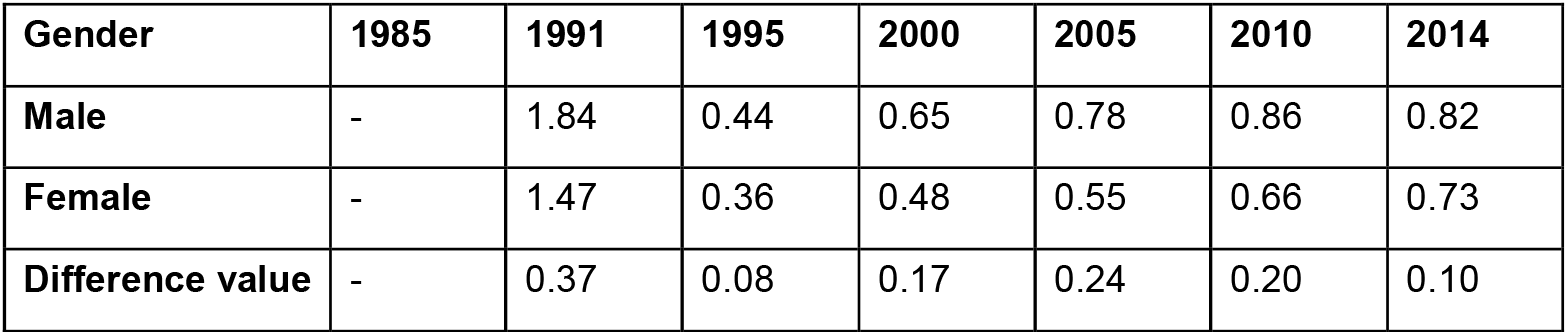
Statistics for the comparison between the link-relative growth rate of the mean height (unit: cm) of boys and girls.

| Gender | 1985 | 1991 | 1995 | 2000 | 2005 | 2010 | 2014 |
| --- | --- | --- | --- | --- | --- | --- | --- |
| Male | - | 1.84 | 0.44 | 0.65 | 0.78 | 0.86 | 0.82 |
| Female | - | 1.47 | 0.36 | 0.48 | 0.55 | 0.66 | 0.73 |
| Difference value | - | 0.37 | 0.08 | 0.17 | 0.24 | 0.20 | 0.10 |

**Table 3.**
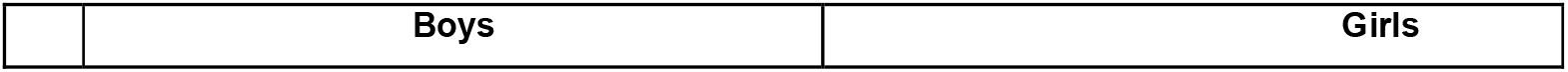
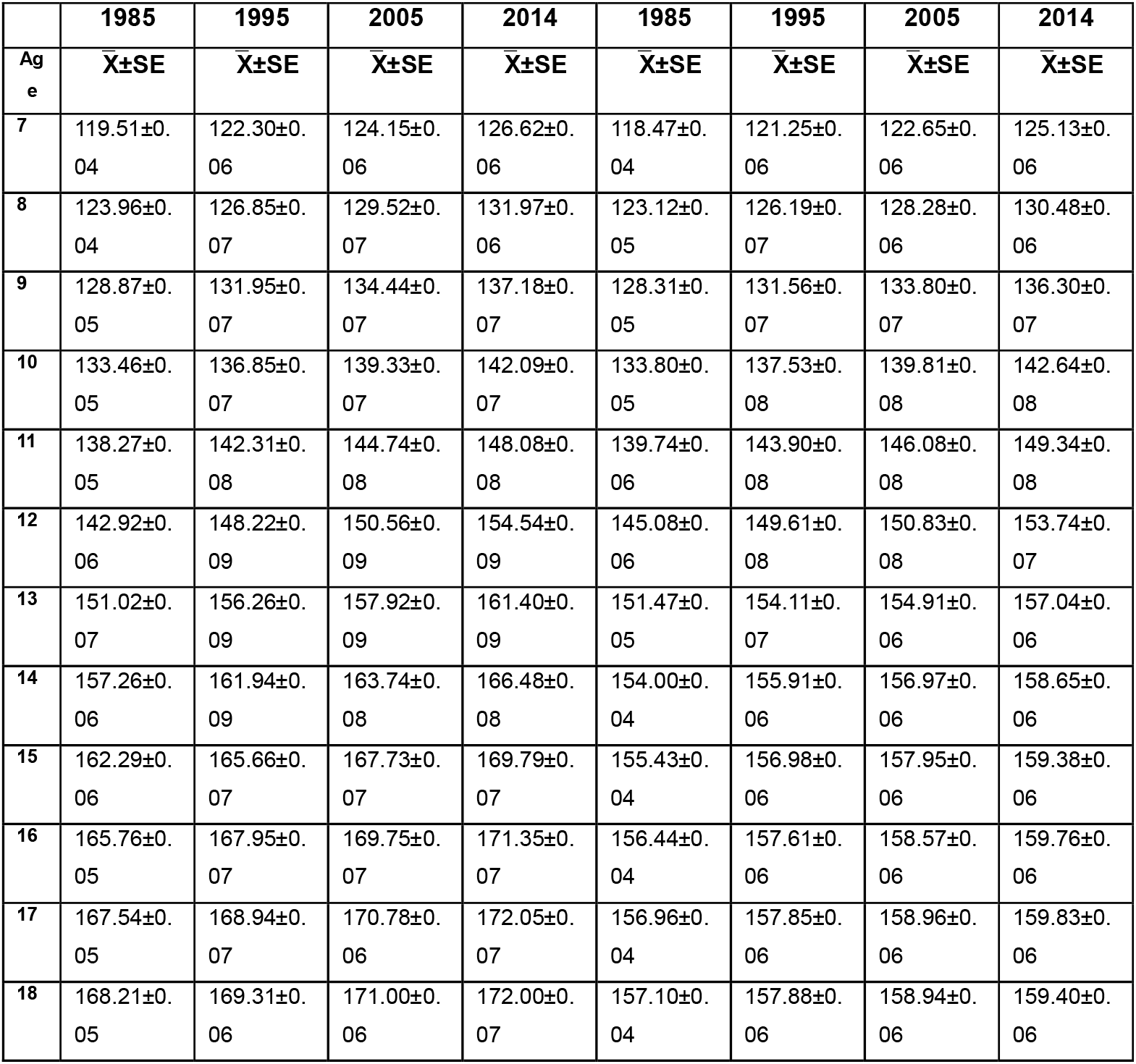
Statistics for the mean height (unit: cm) and standard error of students aged 7-18 in China in 1985, 1995, 2005 and 2014.

|  | 1985 | 1995 | 2005 | 2014 | 1985 | 1995 | 2005 | 2014 |
| --- | --- | --- | --- | --- | --- | --- | --- | --- |
| Age | $\bar{X} \pm SE$ | $\bar{X} \pm SE$ | $\bar{X} \pm SE$ | $\bar{X} \pm SE$ | $\bar{X} \pm SE$ | $\bar{X} \pm SE$ | $\bar{X} \pm SE$ | $\bar{X} \pm SE$ |
| 7 | 119.51±0.04 | 122.30±0.06 | 124.15±0.06 | 126.62±0.06 | 118.47±0.04 | 121.25±0.06 | 122.65±0.06 | 125.13±0.06 |
| 8 | 123.96±0.04 | 126.85±0.07 | 129.52±0.07 | 131.97±0.06 | 123.12±0.05 | 126.19±0.07 | 128.28±0.06 | 130.48±0.06 |
| 9 | 128.87±0.05 | 131.95±0.07 | 134.44±0.07 | 137.18±0.07 | 128.31±0.05 | 131.56±0.07 | 133.80±0.07 | 136.30±0.07 |
| 10 | 133.46±0.05 | 136.85±0.07 | 139.33±0.07 | 142.09±0.07 | 133.80±0.05 | 137.53±0.08 | 139.81±0.08 | 142.64±0.08 |
| 11 | 138.27±0.05 | 142.31±0.08 | 144.74±0.08 | 148.08±0.08 | 139.74±0.06 | 143.90±0.08 | 146.08±0.08 | 149.34±0.08 |
| 12 | 142.92±0.06 | 148.22±0.09 | 150.56±0.09 | 154.54±0.09 | 145.08±0.06 | 149.61±0.08 | 150.83±0.08 | 153.74±0.07 |
| 13 | 151.02±0.07 | 156.26±0.09 | 157.92±0.09 | 161.40±0.09 | 151.47±0.05 | 154.11±0.07 | 154.91±0.06 | 157.04±0.06 |
| 14 | 157.26±0.06 | 161.94±0.09 | 163.74±0.08 | 166.48±0.08 | 154.00±0.04 | 155.91±0.06 | 156.97±0.06 | 158.65±0.06 |
| 15 | 162.29±0.06 | 165.66±0.07 | 167.73±0.07 | 169.79±0.07 | 155.43±0.04 | 156.98±0.06 | 157.95±0.06 | 159.38±0.06 |
| 16 | 165.76±0.05 | 167.95±0.07 | 169.75±0.07 | 171.35±0.07 | 156.44±0.04 | 157.61±0.06 | 158.57±0.06 | 159.76±0.06 |
| 17 | 167.54±0.05 | 168.94±0.07 | 170.78±0.06 | 172.05±0.07 | 156.96±0.04 | 157.85±0.06 | 158.96±0.06 | 159.83±0.06 |
| 18 | 168.21±0.05 | 169.31±0.06 | 171.00±0.06 | 172.00±0.07 | 157.10±0.04 | 157.88±0.06 | 158.94±0.06 | 159.40±0.06 |

In 1984, Tanner called the phenomenon that the Japanese children and adolescents had a height growth rate of over 2.5 cm per 10 years in the period from 1960 to the mid-1980s the “a miracle in the history of human biology” 3. Except Han boys aged 7, 17 and 18, the boys of the other age groups had a height growth rate greater than 2.5 cm per 10 years, from 1985 to 1995, and the mean height growth of the full-range school age group was 2.09 cm (Table 4). In this period, the height growth of the Han boys aged 12 was 5.30 cm and that of the boys aged 13 was 5.24 cm. And the mean height growth of the primary and middle school age group was 3.29 cm. From 2005 to 2014, the height growth of the boys aged 9 to 14 was also greater than 2.5 cm/10 years, and mean height growth of the full-range school age group was 2.49 cm (Table 4). According to an analysis on the mean growth change in the third decade, the mean height growth had a downward trend in the second decade, but the mean height growth increased and did not show a slowdown trend. In the 29 years from 1985 to 2014, the Han boys aged 7-18 increased by 7.61 cm in their average height (Table 4). As can be seen from an analysis on the link-relative growth rate of the mean height of the Han boys, the average growth rate of the boys aged 12 was highest (1.32%) in the 29 years, and that of the boys aged 11 was the second highest (1.15%) (Table 4). The link-relative height growth of the full-range school age group were 2.64 cm, 0.65 cm, 0.96 cm, 0.13 cm, 1.26 cm and 1.23 cm, respectively, in different periods from 1985 to 2014.

**Table 4.** Statistics for F test and T test of the mean height (unit: cm) of boys aged 7-18 in China.

|  | 1985 vs 1995 |  |  |  | 1995 vs 2005 |  |  |  | 2005 vs 2014 |  |  |  | 1985 vs 2014 |  |  |  |
| --- | --- | --- | --- | --- | --- | --- | --- | --- | --- | --- | --- | --- | --- | --- | --- | --- |
| Age | F | Sig. | t'/t | P | F | Sig. | t'/t | P | F | Sig. | t'/t | P | F | Sig. | t'/t | P |
| 7 | 1.13 | < .001 | 36.87 | < .001 | 1.09 | < .001 | 20.94 | < .001 | 1.12 | < .001 | 28.31 | < .001 | 1.10 | < .001 | 95.58 | < .001 |
| 8 | 1.16 | < .001 | 36.10 | < .001 | 1.07 | < .001 | 28.59 | < .001 | 1.13 | < .001 | 26.75 | < .001 | 1.10 | < .001 | 102.73 | < .001 |
| 9 | 1.12 | < .001 | 37.00 | < .001 | 1.03 | > .05 | 26.06 | < .001 | 1.02 | > .05 | 28.81 | < .001 | 1.12 | < .001 | 100.57 | < .001 |
| 10 | 1.25 | < .001 | 38.43 | < .001 | 1.03 | > .05 | 24.47 | < .001 | 1.01 | > .05 | 27.51 | < .001 | 1.23 | < .001 | 99.25 | < .001 |
| 11 | 1.28 | < .001 | 42.05 | < .001 | 1.02 | > .05 | 21.67 | < .001 | 1.06 | < .001 | 29.42 | < .001 | 1.38 | < .001 | 100.33 | < .001 |
| 12 | 1.28 | < .001 | 49.09 | < .001 | 1.03 | > .05 | 19.84 | < .001 | 1.03 | > .05 | 33.96 | < .001 | 1.36 | < .001 | 106.27 | < .001 |
| 13 | 1.07 | < .001 | 45.58 | < .001 | 1.05 | < .05 | 12.58 | < .001 | 1.10 | < .001 | 26.89 | < .001 | 1.02 | > .05 | 92.88 | < .001 |
| 14 | 1.07 | < .001 | 43.06 | < .001 | 1.03 | > .05 | 14.83 | < .001 | 1.17 | < .001 | 23.50 | < .001 | 1.23 | < .001 | 89.18 | < .001 |
| 15 | 1.18 | < .001 | 36.21 | < .001 | 1.06 | < .01 | 20.24 | < .001 | 1.09 | < .001 | 20.41 | < .001 | 1.21 | < .001 | 81.90 | < .001 |
| 16 | 1.02 | > .05 | 26.34 | < .001 | 1.05 | < .05* | 19.02 | < .001 | 1.06 | < .01 | 17.09 | < .001 | 1.01 | > .05 | 68.06 | < .001 |
| 17 | 1.02 | > .05 | 17.62 | < .001 | 1.10 | < .001 | 20.07 | < .001 | 1.03 | > .05 | 13.70 | < .001 | 1.09 | < .001 | 55.81 | < .001 |
| 18 | 1.04 | < .05 | 13.93 | < .001 | 1.10 | < .001 | 18.80 | < .001 | 1.01 | < .001 | 10.85 | < .001 | 1.13 | < .001 | 46.49 | < .001 |

The italicized numeral indicates that the F test was not significant ( $P>0.05$ ), and the Satterthwaite t's test was performed.
And the same below; F means F-Value

In the 29 years from 1985 to 2014, the mean height of the Han girls aged 7-18 in the same age group had a significant growth as times went on (Table 5). The time series analysis was used to make a dynamic analysis on the cumulative growth of the height indexes. In the seven times tests of the girls in the same age group, the girls aged 11 had the largest cumulative growth (9.6 cm); the girls aged 10 had a cumulative growth of 8.84 cm; the girls aged 7 had a cumulative growth of 6.66 cm and a mean height of 125.12 cm; the girls aged 18 had a cumulative growth of 3.32 cm and a mean height of 159.40 cm (Table 5). According to an analysis on the period-to-period growth of the girls, the height growth from 1985 to 1991 is the largest; and the girls aged 9, 10, 11 and 12 had the largest growth (2.57 cm, 2.80 cm and 3.29 cm and 4.62 cm, respectively) (Table 5). According to an analysis on the mean height growth of the full-range school age group in different periods, the mean growths in the seven tests were 2.03 cm, 0.51 cm, 0.69 cm, 0.96 cm, 0.94 cm, and 1.06 cm, respectively (Table 5). This indicates that the height growth of girls decreased slightly from 1991 to 1995, but the height of the Han girls had been increasing steadily from 1995 to 2014.

**Table 5.**
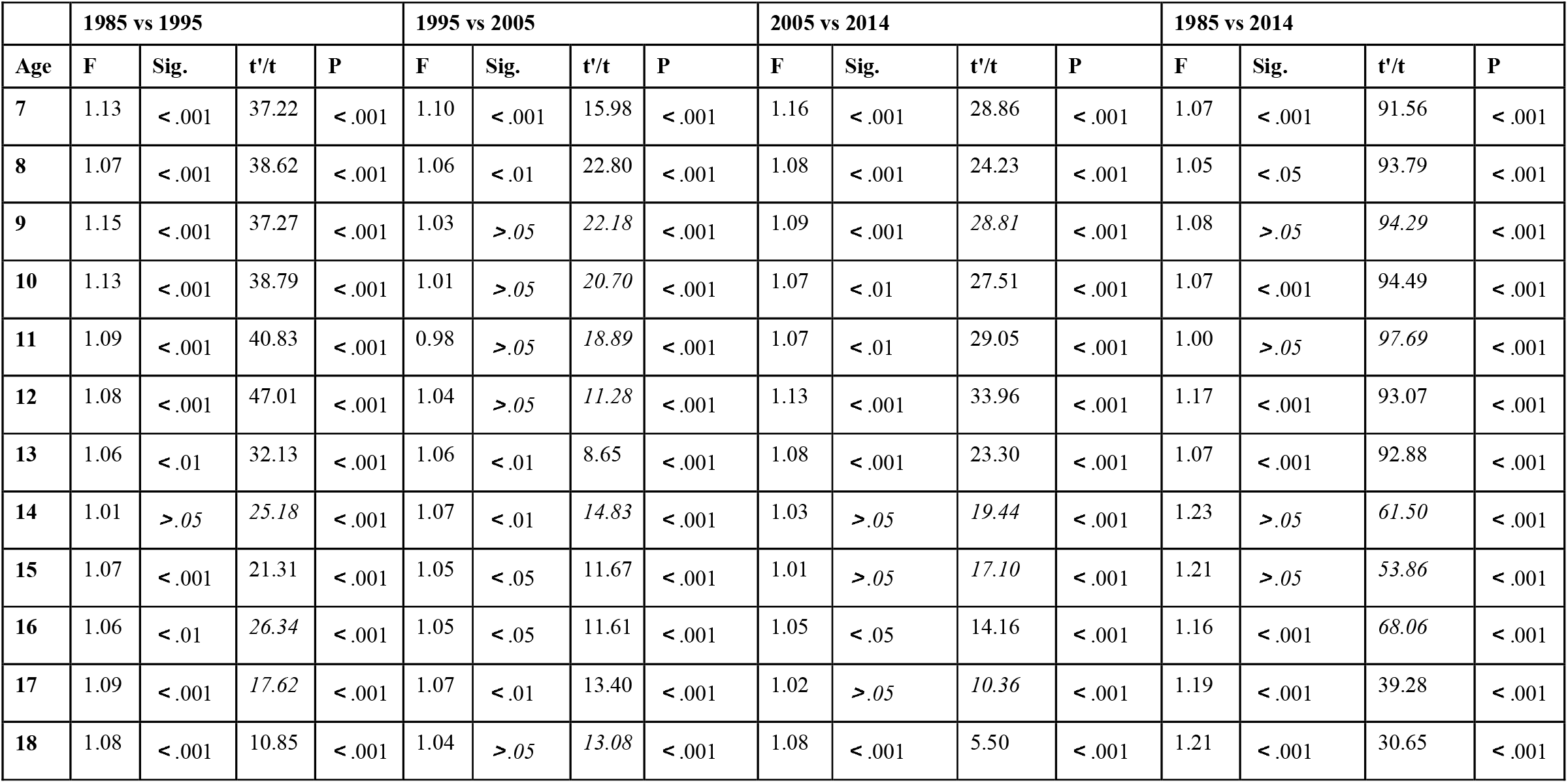
Statistics for F test and T test of the mean height (unit: cm) of girls aged 7-18 in China.

|  | 1985 vs 1995 |  |  |  | 1995 vs 2005 |  |  |  | 2005 vs 2014 |  |  |  | 1985 vs 2014 |  |  |  |
| --- | --- | --- | --- | --- | --- | --- | --- | --- | --- | --- | --- | --- | --- | --- | --- | --- |
| Age | F | Sig. | t'/t | P | F | Sig. | t'/t | P | F | Sig. | t'/t | P | F | Sig. | t'/t | P |
| 7 | 1.13 | < .001 | 37.22 | < .001 | 1.10 | < .001 | 15.98 | < .001 | 1.16 | < .001 | 28.86 | < .001 | 1.07 | < .001 | 91.56 | < .001 |
| 8 | 1.07 | < .001 | 38.62 | < .001 | 1.06 | < .01 | 22.80 | < .001 | 1.08 | < .001 | 24.23 | < .001 | 1.05 | < .05 | 93.79 | < .001 |
| 9 | 1.15 | < .001 | 37.27 | < .001 | 1.03 | > .05 | 22.18 | < .001 | 1.09 | < .001 | 28.81 | < .001 | 1.08 | > .05 | 94.29 | < .001 |
| 10 | 1.13 | < .001 | 38.79 | < .001 | 1.01 | > .05 | 20.70 | < .001 | 1.07 | < .01 | 27.51 | < .001 | 1.07 | < .001 | 94.49 | < .001 |
| 11 | 1.09 | < .001 | 40.83 | < .001 | 0.98 | > .05 | 18.89 | < .001 | 1.07 | < .01 | 29.05 | < .001 | 1.00 | > .05 | 97.69 | < .001 |
| 12 | 1.08 | < .001 | 47.01 | < .001 | 1.04 | > .05 | 11.28 | < .001 | 1.13 | < .001 | 33.96 | < .001 | 1.17 | < .001 | 93.07 | < .001 |
| 13 | 1.06 | < .01 | 32.13 | < .001 | 1.06 | < .01 | 8.65 | < .001 | 1.08 | < .001 | 23.30 | < .001 | 1.07 | < .001 | 92.88 | < .001 |
| 14 | 1.01 | > .05 | 25.18 | < .001 | 1.07 | < .01 | 14.83 | < .001 | 1.03 | > .05 | 19.44 | < .001 | 1.23 | > .05 | 61.50 | < .001 |
| 15 | 1.07 | < .001 | 21.31 | < .001 | 1.05 | < .05 | 11.67 | < .001 | 1.01 | > .05 | 17.10 | < .001 | 1.21 | > .05 | 53.86 | < .001 |
| 16 | 1.06 | < .01 | 26.34 | < .001 | 1.05 | < .05 | 11.61 | < .001 | 1.05 | < .05 | 14.16 | < .001 | 1.16 | < .001 | 68.06 | < .001 |
| 17 | 1.09 | < .001 | 17.62 | < .001 | 1.07 | < .01 | 13.40 | < .001 | 1.02 | > .05 | 10.36 | < .001 | 1.19 | < .001 | 39.28 | < .001 |
| 18 | 1.08 | < .001 | 10.85 | < .001 | 1.04 | > .05 | 13.08 | < .001 | 1.08 | < .001 | 5.50 | < .001 | 1.21 | < .001 | 30.65 | < .001 |

In the periods from 1985 to 1995, 1995 to 2005, and 2005 to 2014, the mean height growth was 2.57 cm, 1.45 cm, and 2.00 cm, respectively (the difference was statistically significant, P<.001** *) (Table 6). In the first decade, the “growth miracle” appeared on girls aged 7 to 13. The “growth miracle” did not appear in the second decade. And in the third decade, the girls aged 9 to 12 had a high growth rate. Their growth were 2.5 cm, 2.83 cm, 3.26 cm and 2.91 cm, respectively. From 1985 to 2014, the mean heights of boys aged 7-18 increased by 6.66 cm, 7.36 cm, 8.00 cm, 8.84 cm, 9.60 cm, 8.66 cm, 5.57 cm, 4.65 cm, 3.95 cm, 3.32 cm, 2.87 cm and 3.32 cm, respectively. Girls aged 11 had the largest growth (9.60 cm). The mean height of girls in the full-range school age group increased by 6.07 cm. The link-relative growth of girls in the full-range school age group was respectively 2.03 cm, 0.51 cm, 0.69 cm, 0.76 cm, 0.94 cm, and 1.06 cm in seven tests. According to an analysis on the period-by-period growth, in the 29 years from 1985 to 2014, girls aged 11 showed the highest average growth rate (1.12%), followed by girls aged 10 and 9, with an average growth rate of 1.07% and 1.01%. The average link-relative growth rate decreased in 1995, while the corresponding figure in the other periods increased as times went on. From 2000 to 2014, the average growth rate was 0.48%, 0.55%, 0.66% and 0.73%, respectively.

**Table 6.**
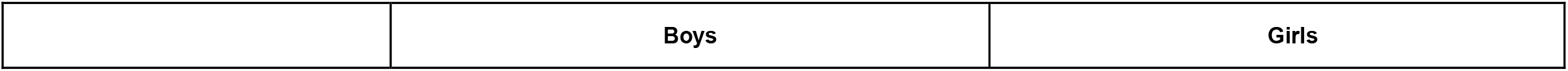
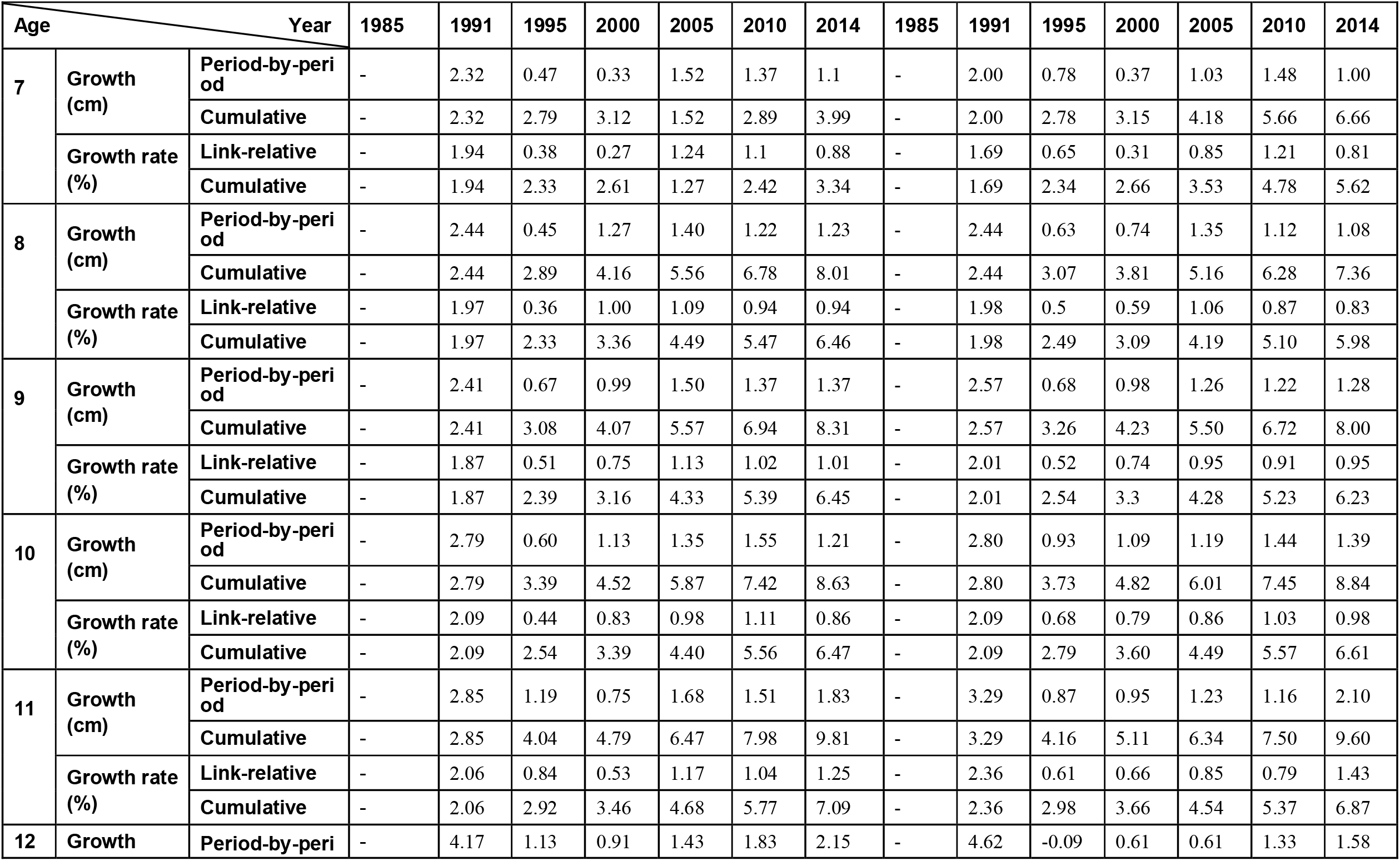
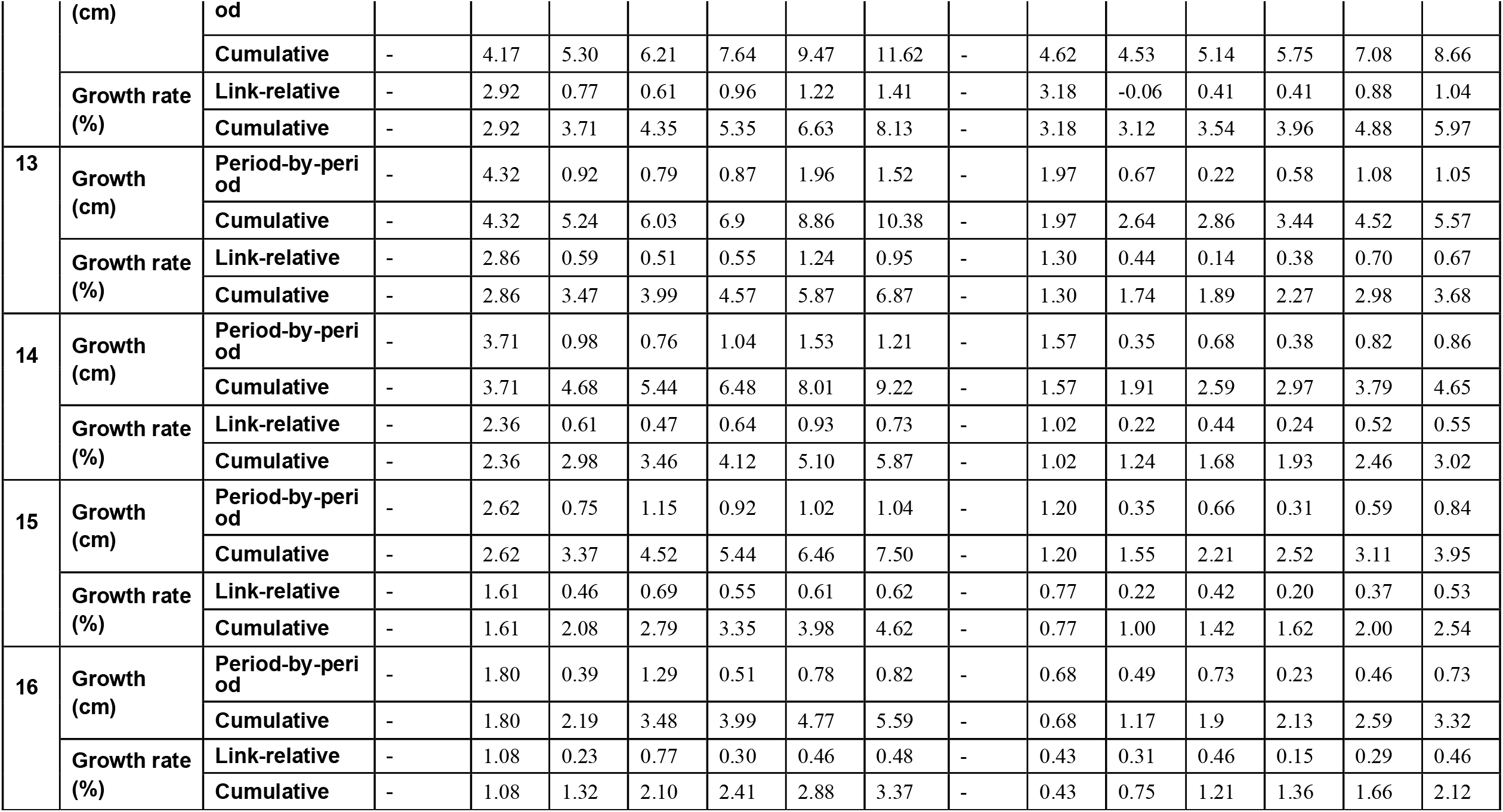
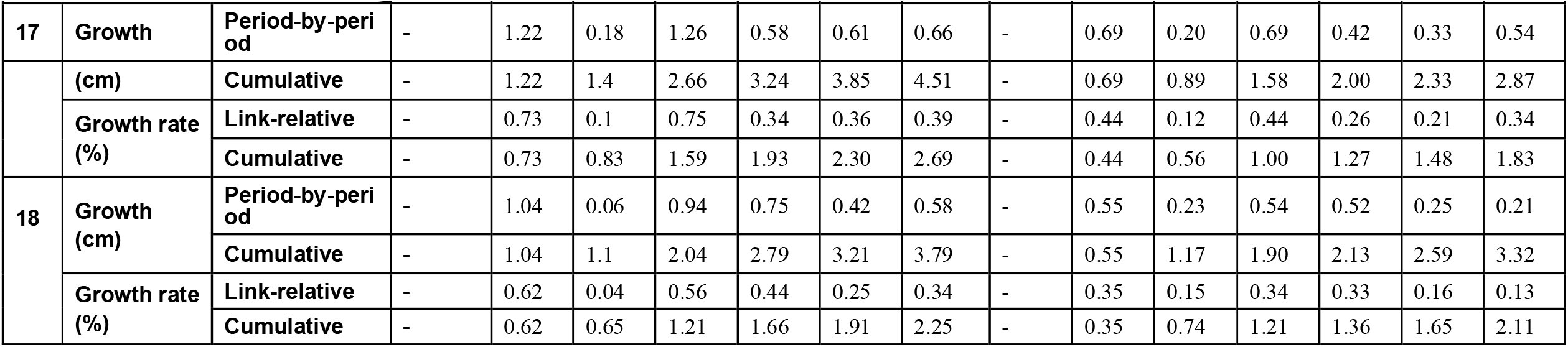
Statistics for the time series growth (unit: cm) and growth rate of the mean height of Han students in China.

| Age \ Year |  |  | 1985 | 1991 | 1995 | 2000 | 2005 | 2010 | 2014 | 1985 | 1991 | 1995 | 2000 | 2005 | 2010 | 2014 |
| --- | --- | --- | --- | --- | --- | --- | --- | --- | --- | --- | --- | --- | --- | --- | --- | --- |
| 7 | Growth (cm) | Period-by-period | - | 2.32 | 0.47 | 0.33 | 1.52 | 1.37 | 1.1 | - | 2.00 | 0.78 | 0.37 | 1.03 | 1.48 | 1.00 |
|  |  | Cumulative | - | 2.32 | 2.79 | 3.12 | 1.52 | 2.89 | 3.99 | - | 2.00 | 2.78 | 3.15 | 4.18 | 5.66 | 6.66 |
|  | Growth rate (%) | Link-relative | - | 1.94 | 0.38 | 0.27 | 1.24 | 1.1 | 0.88 | - | 1.69 | 0.65 | 0.31 | 0.85 | 1.21 | 0.81 |
|  |  | Cumulative | - | 1.94 | 2.33 | 2.61 | 1.27 | 2.42 | 3.34 | - | 1.69 | 2.34 | 2.66 | 3.53 | 4.78 | 5.62 |
| 8 | Growth (cm) | Period-by-period | - | 2.44 | 0.45 | 1.27 | 1.40 | 1.22 | 1.23 | - | 2.44 | 0.63 | 0.74 | 1.35 | 1.12 | 1.08 |
|  |  | Cumulative | - | 2.44 | 2.89 | 4.16 | 5.56 | 6.78 | 8.01 | - | 2.44 | 3.07 | 3.81 | 5.16 | 6.28 | 7.36 |
|  | Growth rate (%) | Link-relative | - | 1.97 | 0.36 | 1.00 | 1.09 | 0.94 | 0.94 | - | 1.98 | 0.5 | 0.59 | 1.06 | 0.87 | 0.83 |
|  |  | Cumulative | - | 1.97 | 2.33 | 3.36 | 4.49 | 5.47 | 6.46 | - | 1.98 | 2.49 | 3.09 | 4.19 | 5.10 | 5.98 |
| 9 | Growth (cm) | Period-by-period | - | 2.41 | 0.67 | 0.99 | 1.50 | 1.37 | 1.37 | - | 2.57 | 0.68 | 0.98 | 1.26 | 1.22 | 1.28 |
|  |  | Cumulative | - | 2.41 | 3.08 | 4.07 | 5.57 | 6.94 | 8.31 | - | 2.57 | 3.26 | 4.23 | 5.50 | 6.72 | 8.00 |
|  | Growth rate (%) | Link-relative | - | 1.87 | 0.51 | 0.75 | 1.13 | 1.02 | 1.01 | - | 2.01 | 0.52 | 0.74 | 0.95 | 0.91 | 0.95 |
|  |  | Cumulative | - | 1.87 | 2.39 | 3.16 | 4.33 | 5.39 | 6.45 | - | 2.01 | 2.54 | 3.3 | 4.28 | 5.23 | 6.23 |
| 10 | Growth (cm) | Period-by-period | - | 2.79 | 0.60 | 1.13 | 1.35 | 1.55 | 1.21 | - | 2.80 | 0.93 | 1.09 | 1.19 | 1.44 | 1.39 |
|  |  | Cumulative | - | 2.79 | 3.39 | 4.52 | 5.87 | 7.42 | 8.63 | - | 2.80 | 3.73 | 4.82 | 6.01 | 7.45 | 8.84 |
|  | Growth rate (%) | Link-relative | - | 2.09 | 0.44 | 0.83 | 0.98 | 1.11 | 0.86 | - | 2.09 | 0.68 | 0.79 | 0.86 | 1.03 | 0.98 |
|  |  | Cumulative | - | 2.09 | 2.54 | 3.39 | 4.40 | 5.56 | 6.47 | - | 2.09 | 2.79 | 3.60 | 4.49 | 5.57 | 6.61 |
| 11 | Growth (cm) | Period-by-period | - | 2.85 | 1.19 | 0.75 | 1.68 | 1.51 | 1.83 | - | 3.29 | 0.87 | 0.95 | 1.23 | 1.16 | 2.10 |
|  |  | Cumulative | - | 2.85 | 4.04 | 4.79 | 6.47 | 7.98 | 9.81 | - | 3.29 | 4.16 | 5.11 | 6.34 | 7.50 | 9.60 |
|  | Growth rate (%) | Link-relative | - | 2.06 | 0.84 | 0.53 | 1.17 | 1.04 | 1.25 | - | 2.36 | 0.61 | 0.66 | 0.85 | 0.79 | 1.43 |
|  |  | Cumulative | - | 2.06 | 2.92 | 3.46 | 4.68 | 5.77 | 7.09 | - | 2.36 | 2.98 | 3.66 | 4.54 | 5.37 | 6.87 |
| 12 | Growth | Period-by-period | - | 4.17 | 1.13 | 0.91 | 1.43 | 1.83 | 2.15 | - | 4.62 | -0.09 | 0.61 | 0.61 | 1.33 | 1.58 |

|  |  |  | Boys |  |  |  |  |  |  | Girls |  |  |  |  |  |  |
| --- | --- | --- | --- | --- | --- | --- | --- | --- | --- | --- | --- | --- | --- | --- | --- | --- |
| Age | Year |  | 1985 | 1991 | 1995 | 2000 | 2005 | 2010 | 2014 | 1985 | 1991 | 1995 | 2000 | 2005 | 2010 | 2014 |
|  | (cm) | od |  |  |  |  |  |  |  |  |  |  |  |  |  |  |
|  |  | Cumulative | - | 4.17 | 5.30 | 6.21 | 7.64 | 9.47 | 11.62 | - | 4.62 | 4.53 | 5.14 | 5.75 | 7.08 | 8.66 |
|  | Growth rate (%) | Link-relative | - | 2.92 | 0.77 | 0.61 | 0.96 | 1.22 | 1.41 | - | 3.18 | -0.06 | 0.41 | 0.41 | 0.88 | 1.04 |
|  |  | Cumulative | - | 2.92 | 3.71 | 4.35 | 5.35 | 6.63 | 8.13 | - | 3.18 | 3.12 | 3.54 | 3.96 | 4.88 | 5.97 |
| 13 | Growth (cm) | Period-by-period | - | 4.32 | 0.92 | 0.79 | 0.87 | 1.96 | 1.52 | - | 1.97 | 0.67 | 0.22 | 0.58 | 1.08 | 1.05 |
|  |  | Cumulative | - | 4.32 | 5.24 | 6.03 | 6.9 | 8.86 | 10.38 | - | 1.97 | 2.64 | 2.86 | 3.44 | 4.52 | 5.57 |
|  | Growth rate (%) | Link-relative | - | 2.86 | 0.59 | 0.51 | 0.55 | 1.24 | 0.95 | - | 1.30 | 0.44 | 0.14 | 0.38 | 0.70 | 0.67 |
|  |  | Cumulative | - | 2.86 | 3.47 | 3.99 | 4.57 | 5.87 | 6.87 | - | 1.30 | 1.74 | 1.89 | 2.27 | 2.98 | 3.68 |
| 14 | Growth (cm) | Period-by-period | - | 3.71 | 0.98 | 0.76 | 1.04 | 1.53 | 1.21 | - | 1.57 | 0.35 | 0.68 | 0.38 | 0.82 | 0.86 |
|  |  | Cumulative | - | 3.71 | 4.68 | 5.44 | 6.48 | 8.01 | 9.22 | - | 1.57 | 1.91 | 2.59 | 2.97 | 3.79 | 4.65 |
|  | Growth rate (%) | Link-relative | - | 2.36 | 0.61 | 0.47 | 0.64 | 0.93 | 0.73 | - | 1.02 | 0.22 | 0.44 | 0.24 | 0.52 | 0.55 |
|  |  | Cumulative | - | 2.36 | 2.98 | 3.46 | 4.12 | 5.10 | 5.87 | - | 1.02 | 1.24 | 1.68 | 1.93 | 2.46 | 3.02 |
| 15 | Growth (cm) | Period-by-period | - | 2.62 | 0.75 | 1.15 | 0.92 | 1.02 | 1.04 | - | 1.20 | 0.35 | 0.66 | 0.31 | 0.59 | 0.84 |
|  |  | Cumulative | - | 2.62 | 3.37 | 4.52 | 5.44 | 6.46 | 7.50 | - | 1.20 | 1.55 | 2.21 | 2.52 | 3.11 | 3.95 |
|  | Growth rate (%) | Link-relative | - | 1.61 | 0.46 | 0.69 | 0.55 | 0.61 | 0.62 | - | 0.77 | 0.22 | 0.42 | 0.20 | 0.37 | 0.53 |
|  |  | Cumulative | - | 1.61 | 2.08 | 2.79 | 3.35 | 3.98 | 4.62 | - | 0.77 | 1.00 | 1.42 | 1.62 | 2.00 | 2.54 |
| 16 | Growth (cm) | Period-by-period | - | 1.80 | 0.39 | 1.29 | 0.51 | 0.78 | 0.82 | - | 0.68 | 0.49 | 0.73 | 0.23 | 0.46 | 0.73 |
|  |  | Cumulative | - | 1.80 | 2.19 | 3.48 | 3.99 | 4.77 | 5.59 | - | 0.68 | 1.17 | 1.9 | 2.13 | 2.59 | 3.32 |
|  | Growth rate (%) | Link-relative | - | 1.08 | 0.23 | 0.77 | 0.30 | 0.46 | 0.48 | - | 0.43 | 0.31 | 0.46 | 0.15 | 0.29 | 0.46 |
|  |  | Cumulative | - | 1.08 | 1.32 | 2.10 | 2.41 | 2.88 | 3.37 | - | 0.43 | 0.75 | 1.21 | 1.36 | 1.66 | 2.12 |
| Age |  | Year | 1985 | 1991 | 1995 | 2000 | 2005 | 2010 | 2014 | 1985 | 1991 | 1995 | 2000 | 2005 | 2010 | 2014 |
| 17 | Growth | Period-by-period | - | 1.22 | 0.18 | 1.26 | 0.58 | 0.61 | 0.66 | - | 0.69 | 0.20 | 0.69 | 0.42 | 0.33 | 0.54 |
|  | (cm) | Cumulative | - | 1.22 | 1.4 | 2.66 | 3.24 | 3.85 | 4.51 | - | 0.69 | 0.89 | 1.58 | 2.00 | 2.33 | 2.87 |
|  | Growth rate (%) | Link-relative | - | 0.73 | 0.1 | 0.75 | 0.34 | 0.36 | 0.39 | - | 0.44 | 0.12 | 0.44 | 0.26 | 0.21 | 0.34 |
|  |  | Cumulative | - | 0.73 | 0.83 | 1.59 | 1.93 | 2.30 | 2.69 | - | 0.44 | 0.56 | 1.00 | 1.27 | 1.48 | 1.83 |
| 18 | Growth (cm) | Period-by-period | - | 1.04 | 0.06 | 0.94 | 0.75 | 0.42 | 0.58 | - | 0.55 | 0.23 | 0.54 | 0.52 | 0.25 | 0.21 |
|  |  | Cumulative | - | 1.04 | 1.1 | 2.04 | 2.79 | 3.21 | 3.79 | - | 0.55 | 1.17 | 1.90 | 2.13 | 2.59 | 3.32 |
|  | Growth rate (%) | Link-relative | - | 0.62 | 0.04 | 0.56 | 0.44 | 0.25 | 0.34 | - | 0.35 | 0.15 | 0.34 | 0.33 | 0.16 | 0.13 |
|  |  | Cumulative | - | 0.62 | 0.65 | 1.21 | 1.66 | 1.91 | 2.25 | - | 0.35 | 0.74 | 1.21 | 1.36 | 1.65 | 2.11 |

### Urban-rural and sex difference in height development

#### Urban-rural differences in height development

According to a comparison between the height of urban and rural boys, the mean height of urban boys aged 7-18 was 4.18 cm greater than that of rural boys in 1985(Table 7). And the urban-rural differences in the mean height of boys were 3.58 cm, 3.18 cm and 2.41 cm, respectively in 1995, 2005 and 2014. It can be seen that the urban-rural difference in the mean height of boys had been gradually narrowed.

**Table 7.** F and t tests of urban-rural differences in the mean height (unit: cm) of Han (boys)

|  | Urban |  | Rural |  | F test |  | t/t' test |  |
| --- | --- | --- | --- | --- | --- | --- | --- | --- |
| Year | $\bar{X} \pm S$ | N | $\bar{X} \pm S$ | N | F-value | P | t'/t | P |
| 1985 | 148.65±18.17 | 102443 | 144.47±18.18 | 102657 | 1.00 | 0.86 | 52.04 | < .001*** |
| 1995 | 151.58±19.45 | 62653 | 148.00±18.16 | 52118 | 1.15 | < .001*** | 6.82 | < .001*** |
| 2005 | 153.62±17.76 | 59233 | 150.44±18.07 | 58447 | 1.03 | < .001*** | 30.45 | < .001*** |
| 2014 | 155.62±17.28 | 53602 | 153.21±17.73 | 53614 | 1.05 | < .001*** | 22.51 | < .001*** |

\* indicates a 5% significance level; \*\* indicates a 1% significance level; \*\*\* indicates a 0.1%significance level, and the same below.

According to a comparison between the height of urban and rural girls (Table 8), the mean height of urban girls aged 7-18 was greater than that of rural boys, and the urban-rural difference in the mean height of girls had been gradually bridged. The results of comparisons showed that the urban-rural differences in the mean height of girls were 3.68 cm, 3.12 cm, 2.62 cm, and 1.98 cm, respectively in 1985, 1995, 2005 and 2014.

**Table 8.**
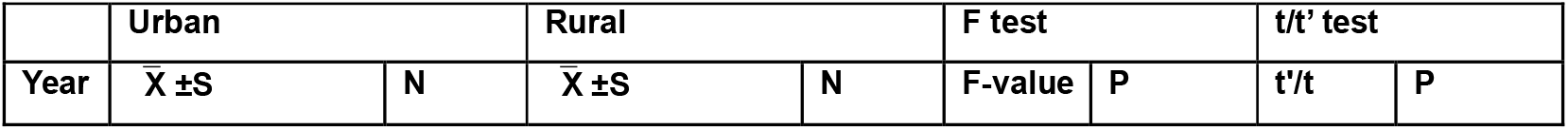
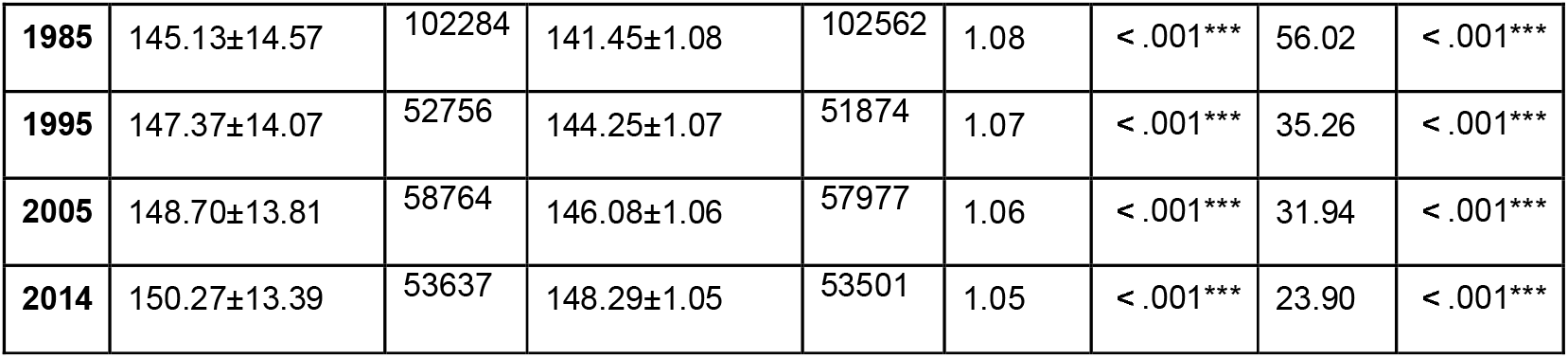
F and t tests of urban-rural differences in the mean height (unit: cm) of Han (girls)

|  | Urban |  | Rural |  | F test |  | t/t' test |  |
| --- | --- | --- | --- | --- | --- | --- | --- | --- |
| Year | $\bar{X} \pm S$ | N | $\bar{X} \pm S$ | N | F-value | P | t'/t | P |
| **1985** | 145.13±14.57 | 102284 | 141.45±1.08 | 102562 | 1.08 | < .001*** | 56.02 | < .001*** |
| **1995** | 147.37±14.07 | 52756 | 144.25±1.07 | 51874 | 1.07 | < .001*** | 35.26 | < .001*** |
| **2005** | 148.70±13.81 | 58764 | 146.08±1.06 | 57977 | 1.06 | < .001*** | 31.94 | < .001*** |
| **2014** | 150.27±13.39 | 53637 | 148.29±1.05 | 53501 | 1.05 | < .001*** | 23.90 | < .001*** |

#### Sex differences in height development

Overall, the sex difference in the mean height of boys and girls aged 7 to 18 had been widened constantly in the period from 1985 to 2014. And the sex differences in 1985, 1995, 2005, and 2014 were 3.27 cm, 4.03 cm, 4.64 cm, and 5.13 cm, respectively (Table 9). According to a comparison between the link-relative height growth rate of boys and girls, the one of boys was higher than that of girls. The sex difference in the height growth rate was the largest in 1991, and the difference was 0.37%. The sex difference in the link-relative growth rate was gradually narrowed from 2005 to 2010 and 2014, and such difference was 0.24%, 0.20% and 0.10%, respectively. According to a comparative analysis on the specific age difference in the height development, girls had the largest height growth at the age of 11, with a mean cumulative growth of 9.60 cm, while boys had the largest height growth at the age of 12, with a mean cumulative growth of 11.62 cm (Table10).

**Table 9.**
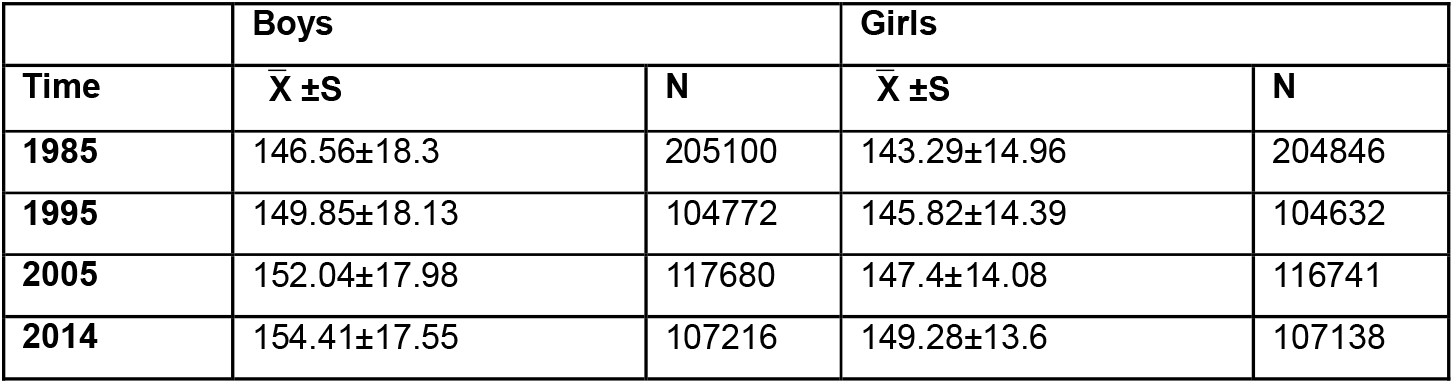
Statistics for the mean value (unit: cm), standard deviation and sample size of the height of the primary and middle school age group.

|  | Boys |  | Girls |  |
| --- | --- | --- | --- | --- |
| Time | $\bar{X} \pm S$ | N | $\bar{X} \pm S$ | N |
| 1985 | 146.56±18.3 | 205100 | 143.29±14.96 | 204846 |
| 1995 | 149.85±18.13 | 104772 | 145.82±14.39 | 104632 |
| 2005 | 152.04±17.98 | 117680 | 147.4±14.08 | 116741 |
| 2014 | 154.41±17.55 | 107216 | 149.28±13.6 | 107138 |

**Table 10.**
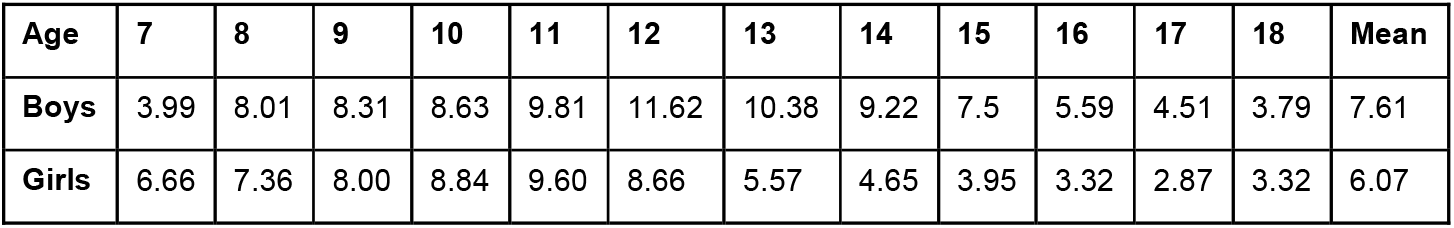
Statistics for comparisons between the mean height growth (unit: cm) of boys and girls.

| Age | 7 | 8 | 9 | 10 | 11 | 12 | 13 | 14 | 15 | 16 | 17 | 18 | Mean |
| --- | --- | --- | --- | --- | --- | --- | --- | --- | --- | --- | --- | --- | --- |
| Boys | 3.99 | 8.01 | 8.31 | 8.63 | 9.81 | 11.62 | 10.38 | 9.22 | 7.5 | 5.59 | 4.51 | 3.79 | 7.61 |
| Girls | 6.66 | 7.36 | 8.00 | 8.84 | 9.60 | 8.66 | 5.57 | 4.65 | 3.95 | 3.32 | 2.87 | 3.32 | 6.07 |

**Table 11.** Model parameter estimation and prediction of the height of Han Boys aged 7-18 in 2024.

| Age | ARIMA Model | AR Coefficient | t-value | P-value | Box-Ljung Test P-value | 2014 Mean | 2024 Predicted value | 95% Confidence interval |
| --- | --- | --- | --- | --- | --- | --- | --- | --- |
| 7 | ( 1,1,0 ) | 0.769 | 2.75 | 0.004 | 0.44 | 126.62 | 128.03↑ | 124.18-131.88 |
| 8 | ( 1,1,0 ) | 0.815 | 3.47 | <0.001 | 0.32 | 131.97 | 133.83↑ | 129.98-137.67 |
| 9 | ( 1,1,0 ) | 0.880 | 5.37 | <0.001 | 0.66 | 137.18 | 139.48↑ | 136.08-142.88 |
| 10 | ( 1,1,0 ) | 0.832 | 3.78 | <0.001 | 0.42 | 142.09 | 143.97↑ | 139.44-148.82 |
| 11 | ( 1,1,0 ) | 0.910 | 7.08 | <0.001 | 0.54 | 148.08 | 151.37↑ | 147.15-155.84 |
| 12 | ( 1,1,0 ) | 0.862 | 4.39 | <0.001 | 0.83 | 154.54 | 158.11↑ | 151.55-165.29 |
| 13 | ( 1,1,0 ) | 0.781 | 2.64 | 0.008 | 0.51 | 161.40 | 163.56↑ | 156.67-171.09 |
| 14 | ( 1,1,0 ) | 0.834 | 3.61 | <0.001 | 0.69 | 166.48 | 168.36↑ | 162.86-174.26 |

|  |  |  |  |  |  |  |  |  |
| --- | --- | --- | --- | --- | --- | --- | --- | --- |
| 15 | ( 1,1,0 ) | 0.865 | 4.60 | <0.001 | 0.30 | 169.79 | 171.49↑ | 167.69-175.48 |
| 16 | ( 1,2,0 ) | -0.908 | 6.88 | <0.001 | 0.31 | 171.35 | 172.98↑ | 171.75-174.23 |
| 17 | ( 1,2,0 ) | -0.866 | 5.13 | <0.001 | 0.09 | 172.05 | 173.34↑ | 172.19-174.50 |
| 18 | ( 2,1,0 ) | 0.714 | 2.56 | 0.01 | 0.14 | 172.00 | 172.86↑ | 171.61-174.13 |

**Table 12.** Model parameter estimation and prediction of the height of Han girls aged 7-18 in 2024.

| A<br>ge | ARIMA<br>Model | AR<br>Coeff<br>icient | t-Val<br>ue | P | MA | t-Val<br>ue | P | Box-Lj<br>ung<br>Test P | 2014<br>Mean | 2023 年<br>Predicted<br>value | 95%<br>CI |
| --- | --- | --- | --- | --- | --- | --- | --- | --- | --- | --- | --- |
| 7 | ( 1,1,0 ) | 0.868 | 4.92 | <0.001 |  | - | <0.001 | 0.39 | 125.13 | 126.78↑ | 123.64-130.09 |
| 8 | ( 1,1,0 ) | 0.848 | 4.13 | <0.001 |  | - | <0.001 | 0.55 | 130.48 | 132.21↑ | 128.38-136.30 |
| 9 | ( 1,1,0 ) | 0.870 | 4.84 | <0.001 |  | - | <0.001 | 0.56 | 136.30 | 138.43↑ | 134.49-142.62 |
| 10 | ( 1,1,0 ) | 0.900 | 6.32 | <0.001 |  | - | <0.001 | 0.62 | 142.64 | 145.08↑ | 141.09-149.30 |
| 11 | ( 1,1,0 ) | 0.862 | 4.39 | <0.001 |  | - | <0.001 | 0.43 | 149.34 | 152.84↑ | 147.48-158.60 |
| 12 | ( 1,1,1 ) | 0.999 | 55.53 | <0.001 | -0.97 | -2.95 | 0.003 | 0.90 | 153.74 | 156.11↑ | 150.50-162.16 |
| 13 | ( 1,1,0 ) | 0.868 | 3.38 | <0.001 |  | - | <0.001 | 0.84 | 157.04 | 158.77↑ | 155.84-161.82 |
| 14 | ( 1,1,0 ) | 0.818 | 3.84 | <0.001 |  | - | <0.001 | 0.09 | 158.65 | 159.95↑ | 157.41-162.57 |
| 15 | ( 1,1,0 ) | 0.854 | 4.28 | <0.001 |  | - | <0.001 | 0.14 | 159.38 | 161.05↑ | 158.76-162.74 |
| 16 | ( 1,1,0 ) | 0.888 | 6.03 | <0.001 |  | - | <0.001 | 0.29 | 159.76 | 161.00↑ | 159.71-162.31 |
| 17 | ( 2,1,0 ) | 0.816 | 3.33 | <0.001 |  | - | <0.001 | 0.08 | 159.83 | 160.72↑ | 159.23-161.32 |
| 18 | ( 1,1,0 ) | 0.840 | 4.63 | <0.001 |  | - | <0.001 | 0.40 | 159.40 | 159.73↑ | 158.79-160.68 |

The research found that the sex difference in the mean height of boys and girls aged 7-18 in the period from 1985 to 2014 increased constantly, and the difference value was 3.27 cm, 4.03 cm, 4.64 cm, and 5.13 cm, respectively in 1985, 1995, 2005 and 2014. That is to say, the sex difference in height development had been constantly widened. This may be because growth potentials of boys and girls are different. Compared with girls, boys are more sensitive to the good living environment in their physical development, but more vulnerable to malnutrition, diseases and adverse social factors 4.Therefore, the sex difference in height can be used as a biological index for measuring whether the living environment is good 10. The height growth trend of Han boys aged 7-18 is more distinct than that of girls, perhaps because the living environment in China has been constantly improved.

### Height prediction

The ARIMA model was used to predict the height mean and 95% confidence interval of Han boys and girls aged 7-18 in 2024. A statistical test was made on the selected model parameter estimates. The P-values of the 7-year-old and 18-year-old models were 0.004 and 0.01, respectively, and the P-values of the other age groups were P<.001. Model residuals were tested by white noise (P>0.05). The 12-year-old girl model was ARIMA (1,1,1); the AR coefficient test P<.001; a‘nd MA coefficient test P=0.003. The P-values of other models were less than 0.001, and the residual sequence was tested by white noise.

In 2024, the mean height of boys and girls in each age group from 7 to 18 will continue to grow. In 2024, boys are expected to have the largest height growth (3.57 cm) at the age of 12, and have the second largest height growth (3.29 cm) at the age of 11. Boys are expected to have a 0.86 cm height growth at the age of 18. Girls are expected to have the largest height growth (3.5 cm) at the age of 11, and have the second largest height growth (2.44 cm) at the age of 10. Girls are expected to have a 0.33 cm height growth at the age of 18.

## Discussion

The Secular Growth Trend is one of the major phenomena in human biology in the last two centuries. It has the following manifestations: Children’s growth level has been improved; pubertal development has been completed at an earlier time; the adult height has increased constantly [11]. In the existing research, Ji Chengye et al. analyzed the secular growth trend of children and adolescents in large cities in China in the period from 1979 to 2005, and found that children and adolescents had the fastest height growth from 1985 to 1995 (the first decade in this study). Meanwhile, the study of Liao Wenke [12] also showed that the height growth of all types of students from 1985 to 1995 was greater than that in the period from 1995 to 2005. This research further calculated the overall trend of height development of Chinese Han students in the third decade. The cumulative growth (2.49 cm) of boy students in the third decade (2005-2014) was higher than that (2.09 cm) in the second decade and lower than that (3.29 cm) in the first decade. Girls showed the same trend of height development. This indicates that Chinese Han students aged 7 to 18 were still in a stage of height development at a high “positive” accelerated growth rate.

From 1985 to 2014, the mean height of the Han boys aged 7-18 in the same age group had increased constantly as times went on. In the 7 tests, the link-relative height, growth of boys in the full-range school age group were 2.64 cm, 0.65 cm, 0.96 cm, 0.13 cm, 1.26 cm and 1.23 cm, respectively in different periods, and the corresponding figure of girls were 2.03 cm, 0.51 cm, 0.69 cm, 0.96 cm, 0.94 cm and 1.06 cm. This indicates that the height growth of students was the largest in the period from 1991 to 1995, perhaps for the following reasons. Firstly, it is related to China’s implementation of reform and opening up in the early 1980s. In 1985, China’s per capita GDP was 855 yuan, and increased to 4, 854 yuan in 1995. The per capita food expenditure of urban areas increased from 352 yuan to 1,766 yuan, and that of rural areas increased from 183 yuan to 768 yuan. Owing to the improvement of the nutritional status and the effective prevention and control of common diseases affecting the health of children and adolescents, the growth and development of children and adolescents had been accelerated for a long time, and the height of children had increased greatly in this period. Secondly, it is also attributed to the rapid development of school sports. At this stage, the national sports facilities of China had an educational orientation. National policies of China gave much attention to the requirements of sports development for sports facilities, kept a watchful eye on students’ constitution and health, and fulfilled the requirements of sports qualification standards for sports facilities [13]. In 1982, China began to implement the “National Physical Training Standard”. This standard broke the old pattern that sports functional departments were in full charge of school sports, created a management model with the participation of various school parties, changed the old evaluation criteria that measured the sports accomplishment solely by their sports skills, and builds a multi-objective and multi-directional evaluation system for measuring the sports accomplishment of students. According to statistics, from 1979 to 1982, the number of people who passed the “National Physical Training Standard” in China was eight times that of 1978. In 1990, Beijing hosted the Asian Games for the first time. The government invested more efforts into the propaganda of sports and improved the sports infrastructure. The enthusiasm of Chinese people for sports had run unprecedentedly high, and the high attention to sports also affected the height development of students. Thirdly, in their growth and development, children and adolescents not only had a secular growth trend, but also overcame the developmental delays caused by the early adverse factors and speeded up to return to the original track of secular growth [14].

According to this research, the sex difference in the mean height of boys and girls aged 7-18 in the period from 1985 to 2014 had increased constantly, and the difference value was 3.27 cm, 4.03 cm, 4.64 cm, and 5.13 cm, respectively in 1985, 1995, 2005 and 2014. That is to say, the sex difference in height development had been constantly widened. This may be because growth potentials of boys and girls are different. Compared with girls, boys are more sensitive to the good living environment in their physical development, but more vulnerable to malnutrition, diseases and adverse social factors. Therefore, the sex difference in height can be used as a biological index for measuring whether the living environment is good or not [10]. The height growth trend of Han boy students aged 7-18 is more distinct than that of girls, perhaps because the living environment in China has been constantly improved.

According to a comparison between the height of urban and rural boys, the mean height of urban boys aged 7-18 was 4.18 cm greater than that of rural boys aged 7-18 in 1985. And the urban-rural differences in the mean height of boys were 3.58 cm, 3.18 cm and 2.41 cm, respectively in 1995, 2005 and 2014. And the urban-rural differences in the mean height of girls were 3.68 cm, 3.12 cm, 2.62 cm, and 1.98 cm, respectively in 1985, 1995, 2005 and 2014. Thus it can be seen that the urban-rural difference in the mean height of primary and middle school students had been gradually bridged. This may be related to reasons for the continuous positive changes in China’s living environment [15].

In 2024, the mean height of boys and girls in each age group from 7 to 18 will continue to grow. In 2024, boys are expected to have the largest height growth (3.57 cm) at the age of 12, while girls are expected to have the largest height growth (3.5 cm) at the age of 11.

## Conclusions

From 1985 to 2014, the mean height of Han students in the same age group from 7 to 18 had increased constantly as times went by. In the period from 1985 to 1991, the height growth of boys/girls was the largest;Sex differences in height development was observed and the gender differences had been widened;We also observed the Urban-rural differences in height development. The mean height of urban students was greater than that of rural students, but their height difference had been narrowed gradually;In 2024, the mean height of boys and girls aged 7-18 are predicted to grow continuously.

## Data Availability

All the data are available from the collection of the books:
1.Student's Constitution and Health Research Group of China. Chinese National Survey on Students' Constitution and Health (CNSSCH)(1985). Beijing: People's Education Press: 1987.
2.Student's Constitution and Health Research Group of China. Chinese National Survey on Students'Constitution and Health (CNSSCH)(1991). Beijing: People's Education Press: 1993.
3.Student's Constitution and Health Research Group of China. Chinese National Survey on Students' Constitution and Health (CNSSCH)(1995). Beijing: People's Education Press: 1996.
4.Student's Constitution and Health Research Group of China. Chinese National Survey on Students'Constitution and Health (CNSSCH)(2000). Beijing: People's Education Press: 2002.
5.Student's Constitution and Health Research Group of China. Chinese National Survey on Students' Constitution and Health (CNSSCH)(2005). Beijing: People's Education Press: 2007.
6.Student's Constitution and Health Research Group of China. Chinese National Survey on Students' Constitution and Health (CNSSCH)(2010). Beijing: People's Education Press: 2012.
7.Student's Constitution and Health Research Group of China. Chinese National Survey on Students' Constitution and Health (CNSSCH)(2014). Beijing: People's Education Press: 2015.

## Author’s Contributions

HCL, WF and ZR were responsible for the conception and design of the study, data collection and the statistical analysis. HCL and WF contributed to interpretation of the findings. The final manuscript was approved by all authors.

## Acknowledgements

We wish to acknowledge the support by the Planning Fund Project of Chinese Ministry of Education under Grant Nos. 20YJA890024.

## Notes

### Competing Interest Statement

The authors have declared no competing interest.

### Author Declarations

I confirm any necessary IRB and/or ethics committee approvals have been obtained.

